# Biases in Attribution Methods for Norovirus and Rotavirus Diarrhea

**DOI:** 10.1101/2025.10.24.25338730

**Authors:** Dehao Chen, Kayoko Shioda, Andrew F. Brouwer, Alicia N. M. Kraay, Andreas Handel, Benjamin A Lopman, Elizabeth T. Rogawski McQuade, Kristin Nelson

## Abstract

**Background:** The estimate of diarrhea burden attributed to a specific enteric pathogen—the population attributable fraction (PAF)—depends on the specific calculation method. Two conventional methods are commonly used to estimate the PAF for enteric infections: the “detection-as-etiology” (DE) method, which defines the PAF as the pathogen prevalence in diarrheal cases; and the “odds-ratio” (OR) method, which expresses the PAF as a function of the OR between pathogen detection and diarrhea. A third, less frequently used method uses the risk ratio (RR) to quantify the strength of infection.

**Methods:** We compared each conventional PAF (DE, OR, or RR PAF) to a model-based (MB) PAF, derived from a transmission model of enteric infection, and defined bias as the crude difference from this “true” MB PAF. We fitted the transmission model to site-specific qPCR data for norovirus and rotavirus detection from MAL-ED (an eight-country birth cohort studying enteric infections) and used the equilibrium states to calculate the MB PAF.

**Results:** For both pathogens, the OR and RR biases were small at all sites (ranging from -5% to +3%), whereas the DE method consistently overestimated the PAF and its bias was the largest of the conventional methods.

**Conclusions:** Our mechanistic model provides an independent alternative to conventional methods, quantifying pathogens-specific enteric burden and the biases in those methods. Our model suggests the DE PAF estimations are consistently biased, and validates the OR and RR methods as feasible, low-bias measures for quantifying enteric burden.

## Introduction

Norovirus and rotavirus are among the leading causes of diarrheal diseases worldwide (1,2): among the enteric viruses, rotavirus and norovirus are the first- and second-leading causes of diarrheal deaths in children under 5 years old globally (1). Attributing disease burden to specific pathogens is critical for guiding the development of targeted interventions, such as vaccines, but accurately assigning diarrhea etiology is challenging in high burden settings where multiple pathogens are frequently detected during diarrhea. The population attributable fraction (PAF) is a measure of the proportion of illness that can be ascribed to a specific cause. There are two commonly used, conventional methods in the literature for characterizing the PAF of diarrhea attributable to a specific enteric pathogen — one is employed by the Foodborne Epidemiology Reference Group (FERG) of WHO (3,4), another is used by the Global Burden of Disease (GBD) study and has been used in large seminal multi-site studies of diarrhea etiology (5–7). However, the PAF estimates from FERG and GBD vary widely in both their values and pathogen burden rankings. In a 2015 study, FERG estimated that 684 million diarrheal cases were attributable to norovirus globally—the largest number among studied enteric pathogens (3). In contrast, in a 2018 study, GBD’s estimate was substantially lower—at 134 million norovirus cases—with most diarrhea episodes were attributed to rotavirus (5). In addition to reflecting differences in study year and data sources, this discrepancy may in part reflect differences in the underlying assumptions of the two methods.

FERG’s method assumes that detecting a pathogen in stool collected during a diarrheal episode implies it is the cause of illness (3). Under this assumption, the prevalence of detection of a pathogen in diarrheal stools estimates the PAF. Hereafter, we refer to this as the “detection-as-etiology” (DE) PAF. The DE PAF has been used to estimate global pathogen-specific diarrhea incidence and mortality, and norovirus prevalence in acute gastroenteritis (1,8,9). However, the presence of a pathogen in diarrheal stool does not necessarily imply the illness is caused by that pathogen. It is not uncommon to detect the pathogen of interest (referred to as the target pathogen hereafter) with other enteric pathogens (coinfection) in diarrheal stools of infants and young children (10). For example, a study from the MAL-ED (Etiology, Risk Factors, and Interactions of Enteric Infections and Malnutrition and the Consequences for Child Health and Development), a birth cohort in low- and middle-income countries evaluating the association of enteric infections with developmental outcomes in children, found that among the norovirus-positive diarrheal stools, 77.7% were also positive for at least one other enteric pathogens (10). A 2008 review suggested coinfections between rotavirus and other enteric pathogens (e.g., astrovirus, *Campylobacter*) were also commonly observed (4). Because the DE method classifies every diarrheal episode in which the target pathogen is detected as etiologic, it may overestimate the PAF by counting all coinfection episodes—including cases where individuals are asymptomatically infected with the target pathogen subsequently symptomatically infected by non-target pathogens—where the target pathogen is not the true cause of diarrhea.

GBD’s method defines the PAF as a function of the odds ratio (OR) for the association between a pathogen’s detection and diarrhea. Hereafter, we refer to this measure as the OR PAF. The OR PAF was first developed for the Global Enteric Multicenter Study (GEMS) and MAL-ED to attribute the etiology of moderate-to-severe diarrhea (MSD) and specific diarrheal syndromes (e.g., overall, acute, and fever-associated diarrhea), respectively (7,11). In addition to MSD, a reanalysis of GEMS estimated pathogen-specific OR PAFs for overall diarrhea (6), and the ORs underlying those OR PAFs from GEMS informed the GBD’s PAF estimate for norovirus diarrhea (5). The validity of this method relies on the assumption that either no confounding exists in the association between pathogen detection and diarrhea, or that any confounding has been fully adjusted for. However, as many enteric pathogens confer only partial immunity after natural infection, re-infection can happen upon recovery. As a result, natural immunity from prior infection may confound the association, leading to a higher proportion of non-diarrheal episodes with asymptomatic infections by the target pathogen than diarrheal episodes (12), resulting in underestimation of OR PAF. Although matching or adjusting for confounders (e.g., age, location, season) can alleviate bias (7), residual confounding (e.g., incompletely measured naturally immunity, or socioeconomic and environmental factors) remains a major challenge in observational studies and may bias OR PAF estimates.

Furthermore, the OR PAF relies on the rare disease assumption such that the OR approximates the risk ratio (RR). Using the RR as the measure of association is the standard method to estimate the PAF and does not require a rare disease assumption, but to our knowledge remains rarely used in studies of enteric infections (6,13–15). Although rarely used, the RR PAF is presumed, in principle, to yield smaller biases than the OR PAF because it does not rely on the rare disease assumption. To evaluate this, we considered the RR PAF as a third empirical method for evaluation in this study.

Mathematical models can simulate the spread of pathogens through a population. Such simulation enables tracking coinfection and immunity from prior infections, which may bias conventional DE, OR, and RR PAF estimates. We developed a mathematical model of the natural history of enteric infection and defined a model-based (MB) PAF to characterize biases in the conventional PAF estimates of diarrhea attributable to norovirus and rotavirus.

## Methods

### Study design of MAL-ED

The study design of MAL-ED has been detailed elsewhere (13,14). Briefly, between 2009 and 2014, routine non-diarrheal stool samples were collected monthly during 24 months of follow-up from birth, at sites of the eight countries: Bangladesh, Brazil, India, Nepal, Pakistan, Peru, South Africa, and Tanzania. During twice-weekly household visits, if a caregiver reported that a child had a diarrheal episode, a diarrheal stool sample was collected for testing. When a twice-weekly visit coincided with a routine monthly visit, the diarrheal stool sample might replace the routine non-diarrheal stool sample (14). Diarrhea incidence was low at sites in Brazil, South Africa, and Tanzania; on average, children experienced <1 episode per person-year in the first 12 months of life. Norovirus and rotavirus were detected using qPCR with the TaqMan Array Card platform (16). A cycle threshold value of 35 was used as the detection limit (16).

### Transmission model

Our mathematical model is a deterministic compartmental model that simulates transmission of a target pathogen in infants and young children. The model follows a susceptible-infected-recovered structure with the possibility of reinfection. The entire population is initially susceptible (*S*) to infection. Susceptible individuals progress to symptomatic infection (*I*_*s*_) under probability *s*_1_ or to asymptomatic infection (*I*_*a*_). Those in *I*_*s*_ recover from symptomatic illness but continue to remain infectious (*I*_*ps*_) without symptoms. After clearing the pathogen (*R*), individuals may be reinfected. The process of reinfection is similar to the initial infection, progressing through symptomatic infection (*I*_*sr*_) under probability s_2_, and post-symptomatic infection (*I*_*psr*_), or asymptomatic reinfection (*I*_*ar*_). The relative infectiousness of asymptomatic infections is *ε* time less than symptomatic infections (Appendix Section 1) (17). We assume a simple birth-death process, with a fixed rate 2, to ensure the model remains demographically dynamic.

Susceptible individuals can develop diarrhea due to non-target (other) pathogens by entering compartment *I*_*other*_ at rate of *λ*_*other*_. Although *λ*_*pathogen*_ can theoretically act along with *λ*_*other*_ in the process of *S* to *I*_*other*_ we simplify the model by assuming individuals experiencing diarrhea from other pathogens cannot be simultaneously susceptible to the target pathogen. The forces of infection (FOI) for the target pathogen (*λ*_*pathogen*_) and for other pathogens (*λ*_*other*_) are defined in Appendix Section 1.

We model coinfection (*I*_*coinf*_) of the target pathogen and other pathogens during diarrhea (4,10). We assume that all coinfections during diarrhea are the result of one of the three processes: 1) individuals in *I*_*other*_ acquire the target pathogen and transition to *I*_*coinf*_, yielding diarrheal episodes in which the target pathogen is present but not the sole pathogen detected, 2) individuals already infected with the target pathogen symptomatically (*I*_*s*_, *I*_*sr*_) subsequently infected by a non-target pathogen and enter *I*_*coinf*_, and 3) individuals already infected with the target pathogen asymptomatically (*I*_*ps*_, *I*_*a*_, *I*_*psr*_, *I*_*ar*_) subsequently infected by a non-target pathogen and enter *I*_*coinf*_. Because norovirus and rotavirus generally have shorter or similar diarrhea durations than other enteric pathogens (Table S1), we assume coinfection ends upon recovery from the non-target pathogens, which coincides with recovery from the target pathogen. The model’s ordinary differential equations are provided in Appendix Section 1.

We employ the same model structure but different parameter values for norovirus and rotavirus. For each target pathogen, we use empirical infection durations estimated from publicly available MAL-ED data or from the literature (see Table S1). We assume recovery rates for infections to be the inverse of infections durations, following an exponential distribution.

### Estimation of transmission model parameters

We defined the model’s likelihood as the joint probability of three observed endpoints from stool samples of infants ≤ 12 months of age in the MAL-ED cohort: 1) annual incidence of all-cause diarrhea, 2) cumulative prevalence of the target pathogen (norovirus GII [referred to as norovirus] or rotavirus) in diarrheal (symptomatic) stool samples, and 3) cumulative prevalence of the target pathogen in non-diarrheal (asymptomatic) stool samples (Appendix Section 2).

Although MAL-ED followed participants from birth to 24 months, we used a simpler approach and focused on a single age group—0-12 months—which has the highest diarrheal burden (16). We chose norovirus GII for analysis over norovirus GI, as norovirus GII is more associated with person-to-person transmission (18). We applied sample-importance resampling to infer the site-specific model parameters—transmission coefficients of the target pathogen (*β*_*pathogen*_) and other pathogens (*β*_*other*_) and the probabilities of primary (*s*_1_) and secondary (*s*_2_) symptomatic infection. First, we drew 10,000 sets of Sobol samples of (*s*_1_, *s*_2_) from their uniform priors on [0,1]. For each draw *i*, we estimated the negative log-likelihood (NLL_i_) at the maximum-likelihood estimates (MLE) of *β*_*pathogen*_ and *β*_*other*_. We converted NLL_i_ to a probability using 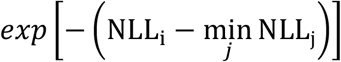, where the minimum was taken over all values of *j* (*j* = 1,2,3, …,10,000). Using the probability, we resampled 10,000 parameter sets with replacement to approximate the joint posterior of the four parameters. For each resampled parameter set *j*, we solved for the model’s equilibrium compartment sizes to derive the corresponding epidemiologic parameters (e.g., PAFs and their biases, coinfection prevalence). We summarized each estimate by its posterior median and 95% confidence interval (CI, 2.5^th^ and 97.5^th^ percentiles) of the resampled posterior distribution. This procedure enabled quantification of uncertainty in the model parameters and in their resulting epidemiologic parameters. We compared the simulated endpoints defining the likelihood to their observed values to assess the model’s goodness-of-fit.

### Population attributable fractions

We used the relative sizes of the compartments at equilibrium (labeled with superscript ^*^ thereafter) to reproduce the DE and OR PAF estimates and to derive a “true” MB PAF. Since the DE PAF is the fraction of diarrhea episodes in which the target pathogen is present (*I*^*\**^_*s*_, *I*^*\**^_*sr*_, and *I*^*\**^_*coinf*_) among all diarrhea episodes (*I*^*\**^_*s*_, *I*^*\**^_*sr*_, or *I*^*\**^_*coinf*_, and *I*^*\**^_*other*_), we formulated the equilibrium DE PAF as follows:

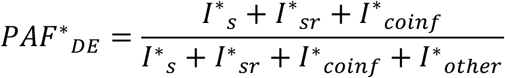

Using the model’s equilibrium sizes, we classified the compartments—as they would be in an observational study—into cases (symptomatic population) and non-cases (asymptomatic population) by exposure status (presence versus absence of the target pathogen, i.e., exposed versus unexposed). This classification was color-coded in **Figure 1**. Among cases, the exposed individuals were those in *I*^*\**^_*s*_, *I*^*\**^_*sr*_, or *I*^*\**^_*coinf*_, while the unexposed were those in *I*^*\**^_*other*_. Because the target pathogen was present in the symptomatic *I*^*\**^_*coinf*_, we classified individuals in that compartment as exposed cases. Among non-cases, the exposed individuals were those in *I*^*\**^_*a*_, *I*^*\**^_*ar*_, *I*^*\**^_*ps*_, and *I*^*\**^_*psr*_, while the unexposed were those in *s*^*\**^ and *R*^*\**^. Hence, the equilibrium OR is:

**Figure 1.**
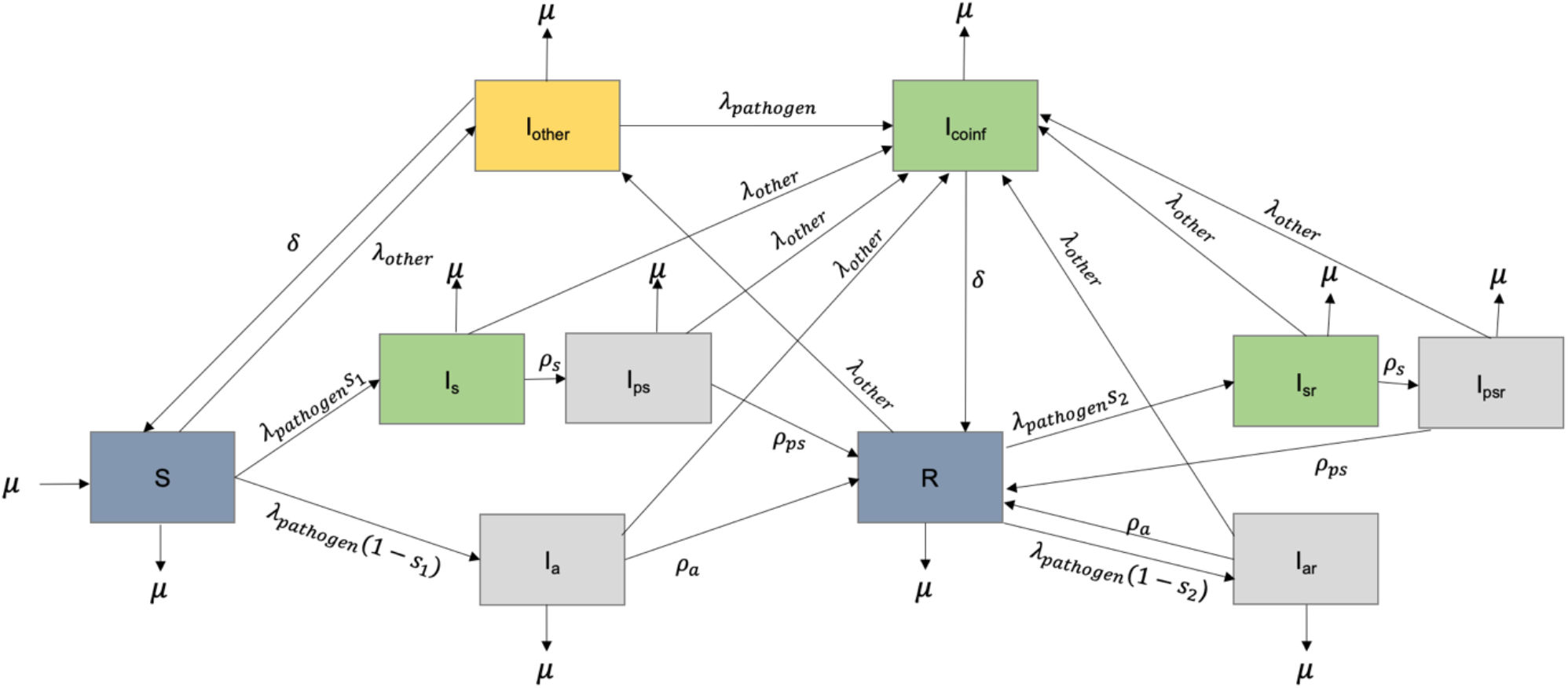
Compartmental model of the natural history of enteric infection. The population is susceptible to symptomatic (*I*_*s*_) or asymptomatic (*I*_1_) infection by a target pathogen. After symptomatic infection, there is a period of post-symptomatic infection. Upon recovery, individuals can be reinfected similar to the initial infections. The compartments are color-coded to represent their classifications for calculating the odds of infection given diarrhea: exposed cases (green), exposed non-cases (grey), unexposed cases (yellow), and unexposed non-cases (blue). Model parameters and ordinary differential equations are shown in the Online Appendix.

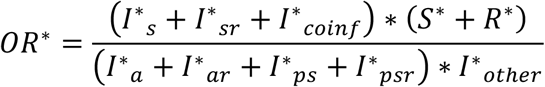

Following the standard formulation (5,7,11), the equilibrium OR PAF is:

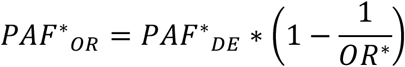

We applied the same exposure-disease classification to derive the RR for the association between pathogen infection and diarrhea risk. At equilibrium, the risk of diarrhea among those infected with the target pathogen (exposed) is (*I*^*\**^_*s*_ + *I*^*\**^_*sr*_ + *I*^*\**^_*coinf*_)/(*I*^*\**^_*s*_ + *I*^*\**^_*sr*_ + *I*^*\**^_*coinf*_ + *I*^*\**^_*a*_ + *I*^*\**^_*ar*_ + *I*^*\**^_*ps*_ + *I*^*\**^_*psr*_), and the risk among those not infected by the target pathogen (unexposed) is (*I*^*\**^_*other*_)/(*I*^*\**^_*other*_ + *s*^*\**^ + *R*^*\**^). The equilibrium RR is:

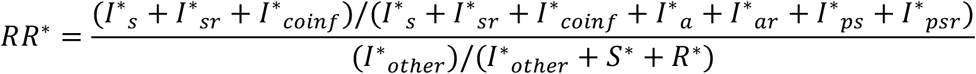

Following the standard formulation (19), the equilibrium RR PAF is:

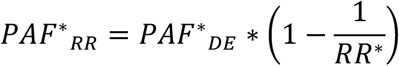

We defined the MB PAF as the “true” fraction of diarrheal episodes (*I*^*\**^_*s*_ + *I*^*\**^_*sr*_ + *I*^*\**^_*coinf*_ + *I*^*\**^_*other*_) attributable to the target pathogen. Primary (*I*^*\**^_*s*_) and secondary (*I*^*\**^_*sr*_) symptomatic cases were fully attributable to the target pathogen. For coinfection, we allocated attribution based on the seven infection pathways entering *I*^*\**^_*coinf*_: one pathway via the FOI of the target pathogen (*λ*_*target*_), which was the only contributor to attribution of the target pathogen; six pathways via the FOI of the other pathogens (*λ*_*other*_). So, the weight on the target pathogen for *I*^*\**^_*coinf*_ was:

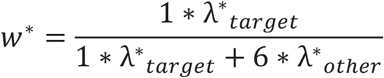

Hence, at equilibrium, the MB PAF combines fully attributable symptomatic cases and a subset of coinfections:

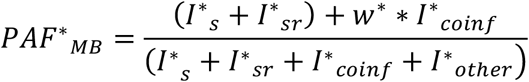

We quantified biases of each conventional PAF (DE, OR, or RR PAF) as the crude difference between its value and the “true” MB PAF calculated at model equilibrium. We expressed the crude difference as a percentage (e.g., 0.2 = 20%). We described the DE, OR, and RR PAF were overestimated (overestimation) when they were larger than the MB PAF and were underestimated (underestimation) they were lower than the MB PAF. To understand the associations between three key epidemiologic outputs (i.e., all-cause diarrhea incidence, coinfection prevalence in diarrheal cases, and asymptomatic infection prevalence) and 1) the biases, and 2) the MB PAF, we calculated the Pearson correlation coefficient for each association. Analyses were performed in R (20), and the scripts were available at: https://github.com/lopmanlab/enteric_paf.

## Results

### Model-estimated epidemiologic measures

Figure 2 compares the simulated and observed endpoints, defining the likelihood, from the norovirus and rotavirus models: 1) pathogen detection prevalence in symptomatic samples, 2) pathogen detection prevalence in asymptomatic samples, 3) incidence rate of all-cause diarrhea per person-year. Although not defined in the likelihood, we compared the simulated ORs to their observed values in Figure S1. In all settings, the simulated estimates of the endpoints and the ORs closely matched the observed values, indicating adequate goodness-of-fit of our models. The incidence rate of all-cause diarrhea was low—less than one episode per person-year—in Brazil, South Africa, and Tanzania. Because the model failed to reach equilibrium despite a prolonged simulation for 100 years, we did not report the equilibrium estimates for norovirus in Brazil, resulting from the low observed number of norovirus detections among diarrhea episodes at this site.

**Figure 2.**
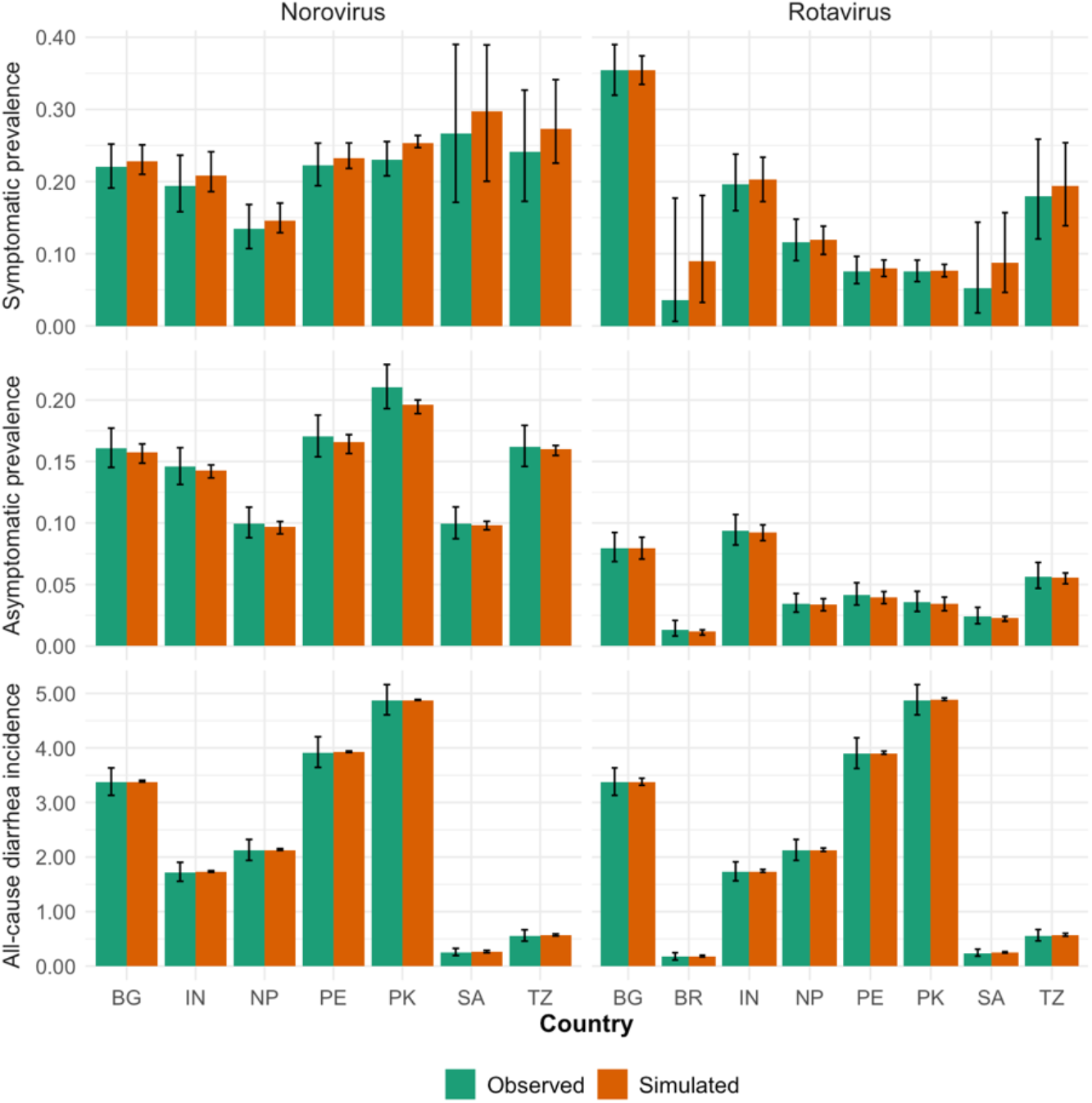
Simulated endpoints used in the likelihood definition for the norovirus and rotavirus transmission models, with corresponding observed data. The endpoints include site-specific symptomatic and asymptomatic cumulative prevalence of pathogen detection and all-cause diarrhea incidence rate (episodes per person-year). Observed data are plotted with Wald-based 95% confidence intervals. Sites are labeled by country abbreviation: Bangladesh (BG), Brazil (BR), India (IN), Nepal (NP), Pakistan (PK), Peru (PE), South Africa (SA), and Tanzania (TZ).

The simulated prevalence of symptomatic coinfection was higher for norovirus than for rotavirus, and the ORs for the association between pathogen detection and diarrhea were closer to the null value of one for norovirus than rotavirus at all sites (Figure S2).

The estimated primary (*s*_1_) and secondary (*s*_2_) symptomatic infection probabilities were generally higher for rotavirus than for norovirus for all countries, except for *s*_1_ in South Africa (Figure S3). Except in South Africa and Tanzania in the norovirus model where *s*_1_ was marginally higher than *s*_2_, *s*_1_ remains lower than _2_ in all settings. The fitted transmission coefficients of the target pathogen (*β*_*pathogen*_) and other pathogens (*β*_*other*_) were included in Figure S3.

### Population attributable fractions

Figure 3 shows the country-specific PAF estimates for norovirus and rotavirus by method. In all countries, the DE PAF was the largest. While the OR PAF was slightly larger than RR PAF for both pathogens, their country-to-country rank orderings were identical. The MB PAF fell between the DE and OR PAFs in South Africa and Tanzania for norovirus and in Tanzania for rotavirus. The OR PAF was higher than the MB PAF in Bangladesh, Nepal, Pakistan, and Peru for norovirus, and in all countries except Tanzania for rotavirus. Of the MB PAF’s associations with the three key epidemiologic outputs, only the correlation with all-cause diarrhea incidence was significant, and only for norovirus (r = -0.81, p value <0.05) — at sites where all-cause diarrhea incidence was higher, the MB PAF was closer to zero (Table S2).

**Figure 3.**
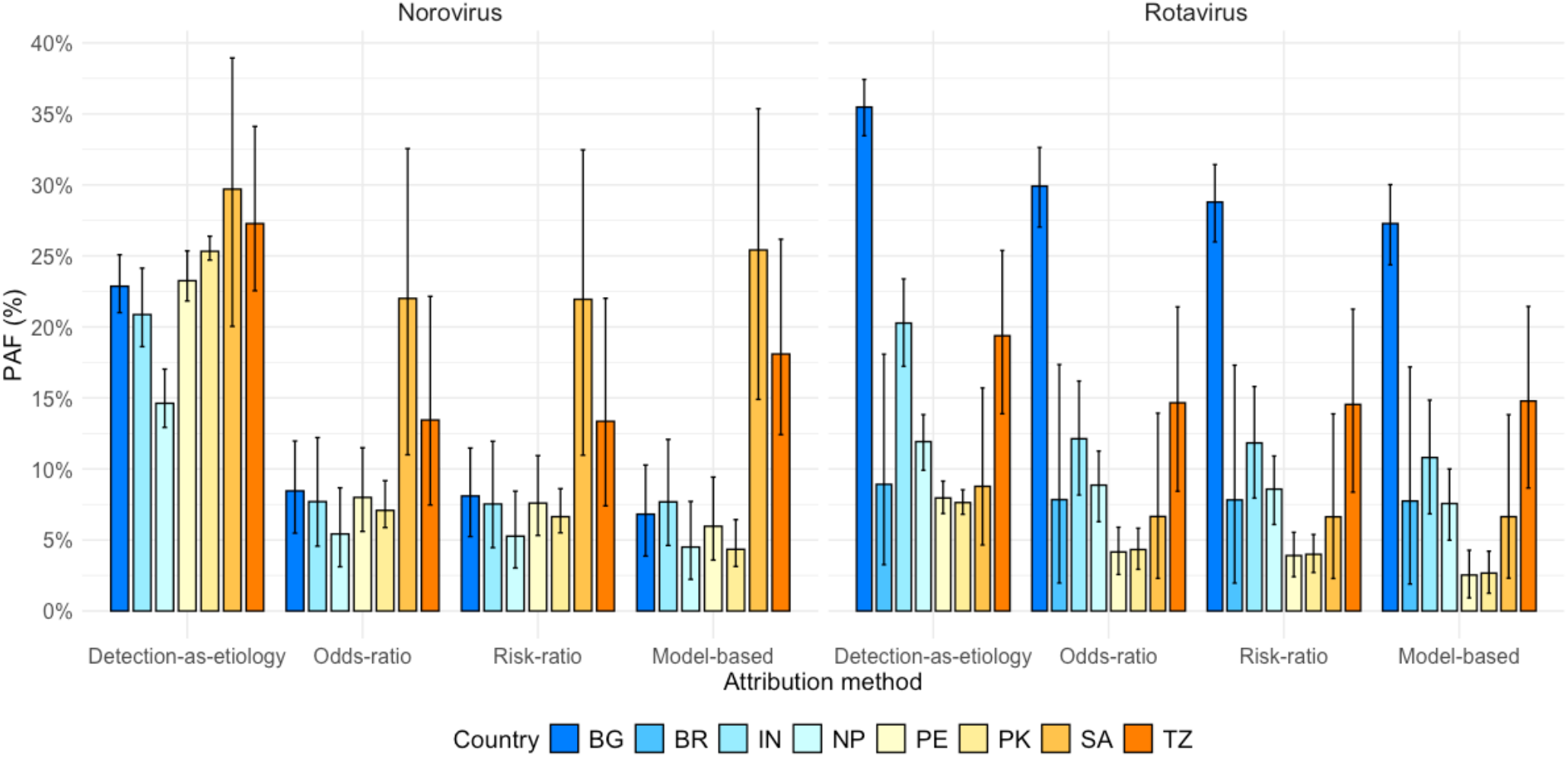
Model-realized site-specific population attributable fractions (PAFs) for norovirus and rotavirus diarrhea. Sites are labeled by country abbreviation: Bangladesh (BG), Brazil (BR), India (IN), Nepal (NP), Pakistan (PK), Peru (PE), South Africa (SA), Tanzania (TZ).

### Biases in population attributable fractions

Figure 4 presents the bias for the DE, OR, and RR methods for norovirus and rotavirus diarrhea across sites. For both pathogens, biases in OR and RR PAFs were small at all sites (-5% to +3%). The PAF was overestimated by the DE method, which has the largest bias across sites among the conventional methods. The magnitude of the bias in the DE PAF for norovirus (4% to 21%) was greater than that for rotavirus (1% to 9%) across sites. From site to site, the bias in OR PAF was similar in magnitude to the bias in RR PAF —though at sites where the PAF was overestimated by the RR and OR methods, the bias in OR PAF slightly exceeded that in the RR PAF; at sites where the PAF was underestimated by the RR and OR PAFs, the bias in RR PAF was larger than the bias in OR PAF in absolute value.

**Figure 4.**
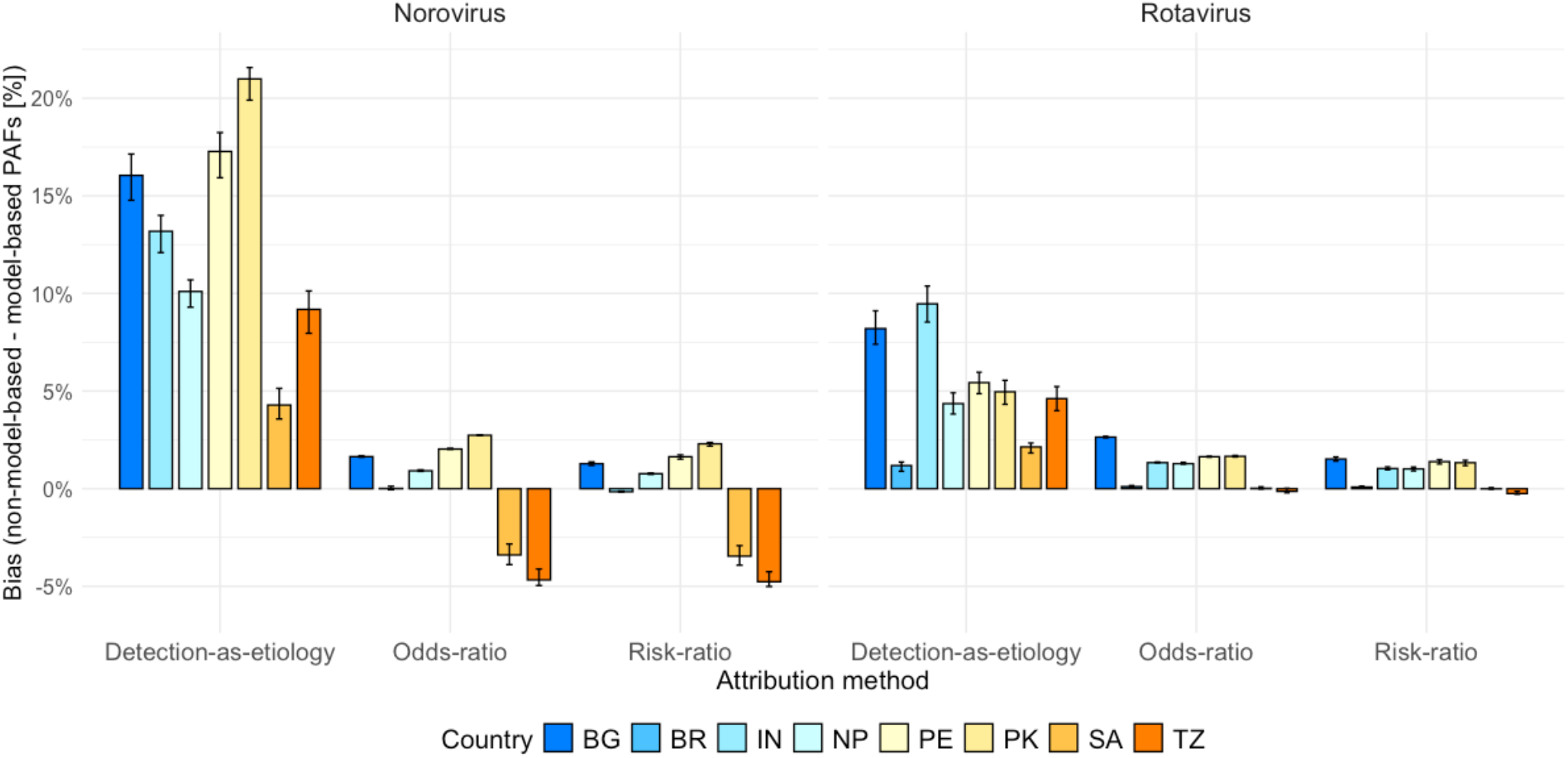
Model-realized site-specific biases of conventional population attributable fraction (PAF) methods for norovirus and rotavirus diarrhea. The bias of each conventional PAF is quantified as the crude difference between that PAF and the MB PAF, expressed as a percentage (e.g., 0.2 = 20%). Sites are labeled by country abbreviation: Bangladesh (BG), Brazil (BR), India (IN), Nepal (NP), Pakistan (PK), Peru (PE), South Africa (SA), Tanzania (TZ).

### Correlation between bias and epidemiologic outputs

Table 1 presents the correlations between each bias in the attribution methods and the three key epidemiologic outputs—annual incidence of all-cause diarrhea, symptomatic coinfection prevalence, and asymptomatic infection prevalence—from the norovirus and rotavirus models. The PAF was significantly overestimated by the DE method when the all-cause diarrheal incidence, coinfection prevalence, and asymptomatic infection for norovirus were high, and when coinfection and asymptomatic infection for rotavirus were high. The PAF was significantly overestimated by the RR and OR methods when the all-cause diarrheal incidence was high.

**Table 1.** Correlations between biases of population attributable fraction methods and key epidemiologic outputs from the norovirus and rotavirus transmission models.

| Attribution Method | All-cause diarrhea |  | Coinfection |  | Asymptomatic |  |
| --- | --- | --- | --- | --- | --- | --- |
|  | incidence |  | prevalence in |  | infection prevalence |  |
|  |  |  | diarrheal cases |  |  |  |
|  | Norovirus | Rotavirus | Norovirus | Rotavirus | Norovirus | Rotavirus |
| Detection as | 0.95 | 0.47 | 0.92 | 0.98 | 0.83 | 0.95 |
| etiology | (<0.05)<sup>&</sup> | (0.24) | (<0.05) | (<0.05) | (<0.05) | (<0.05) |
| Odds ratio | 0.92 | 0.84 | 0.57 | 0.56 | 0.40 | 0.51 |
|  | (<0.05) | (<0.05) | (0.19) | (0.15) | (0.38) | (0.20) |
| Risk ratio | 0.91 | 0.89 | 0.55 | 0.48 | 0.37 | 0.40 |
|  | (<0.05) | (<0.05) | (0.21) | (0.23) | (0.41) | (0.32) |

<sup>&</sup>Pearson correlation (p value)

## Discussion

Compared with PAF estimates from a mechanistic model of enteric infection, conventional methods (DE, OR, and RR) exhibited varying degrees of bias in quantifying pathogen-specific diarrhea burden. The DE method consistently overestimated the PAF across all MAL-ED sites, with a larger bias for norovirus than for rotavirus (4% to 21% for norovirus and 1% to 9% for rotavirus). In contrast, OR and RR PAFs exhibited smaller biases across sites—-5% to +3%—for norovirus and rotavirus. Given the minimal bias in the OR and RR methods, we expect them to be sufficiently accurate for PAF estimation and therefore recommend them over the DE method.

Despite their low magnitude, we did find biases for the OR and RR PAFs, and both biases were statistically significantly correlated with diarrheal incidence. Underestimation of PAF by the OR and RR methods tended to occur in countries with low diarrhea incidence—e.g., South Africa and Tanzania, where, on average, each child had fewer than one diarrheal episode during the first 12 months of life (Figure 2). A likely explanation is natural immunity from prior infection: children with prior infection are protected against diarrhea but may still shed the pathogen without symptoms (asymptomatic infection). This inflates pathogen prevalence in controls relative to cases, and biasing OR/RR toward the null, thereby lowering the PAF. Also, the effect of natural immunity at these sites with low diarrhea incidence is reflected by the relatively low probability of secondary symptomatic infection (*s*_2_). By contrast, at sites with higher diarrheal incidence, the OR and RR methods overestimated the PAF, and the overestimation in OR PAF exceeded that in the RR PAF; this result might suggest a greater violation of the rare-disease assumption for OR under high diarrheal incidence. Still, at such high incidence, these overestimations were small (maximum at +3%), which suggested estimates from the OR and RR methods may be sufficiently reliable for public health planning, even in high-burden settings.

A consistent overestimation of PAF by DE is expected because DE is defined primarily by symptomatic coinfection prevalence. As shown in Table 1, when coinfection prevalence is higher, more overestimation by DE exists. Similarly, the overestimation by DE under high diarrheal incidence in the norovirus model—but not in the rotavirus model—likely reflects the higher extent of coinfection for norovirus, which drives up all-cause diarrhea incidence. The statistically significant correlation between asymptomatic infection prevalence and DE bias may also be explained by coinfection: as asymptomatic infection from the target pathogen increases, other (non-target) pathogens are more commonly the primary cause of diarrhea, allowing individuals already asymptomatically infected to acquire secondary symptomatic infections that manifest as coinfections during diarrheal episodes.

This study has several limitations. First, our model assumed that the target pathogen and other pathogens had independent effects on whether infection was symptomatic or not. However, synergistic effects between rotavirus and coinfecting pathogens have been documented (21). If incorporated into the model, such synergy may increase each pathogen’s diarrheal potential, yielding more cases fully attributable to rotavirus and a higher share of coinfection allocated to it. Hence, the rotavirus MB PAF may be inflated, which could in turn affect the magnitude of bias estimated using the MB PAF. Second, because our objective was to compare the PAF of the conventional methods to the MB PAF, the model compartments did not explicitly include maternally derived immunity. This exclusion may confound our estimates of primary symptomatic infection probability (*s*_1_). Indeed, *s*_1_ was estimated to be lower than the secondary symptomatic infection probability (*s*_2_) in many settings (Figure S3). If we explicitly modeled maternal immunity, we would expect s_1_ to remain consistently higher than *s*_2_, since *s*_2_ reflects immunity acquired from primary infection after maternal immunity wanes. Consequently, we could not characterize acquired immunity as the decline from *s*_1_ to *s*_2_. However, we do not expect this misspecification to affect the results, as the effect of maternal immunity is already absorbed into *s*_1_. Third, unlike established literature using OR PAF for attribution of enteric infections, where OR PAFs rest on ORs adjusted for covariates (e.g., infection by other pathogens, sociodemographic confounders) and temporal dependency (5,7,22), the ORs simulated from our model was crude and was directly calculated from counts of diarrheal and non-diarrheal episodes by exposure status aggregated from all follow-ups. Further, we did not incorporate pathogen quantity when characterizing the model-realized ORs; by contrast, pathogen quantity was incorporated when calculating OR PAFs in the GEMS and MAL-ED studies (6,16).

As an alternative to conventional methods, our mechanistic model provides an independent measure for enteric burden that quantifies the biases in conventional estimates. Fitted to MAL-ED data where diarrhea incidence tends to be high, our model suggests the DE method overestimates the PAF, whereas the OR and RR methods likely provide low-bias measures for quantifying the relative burden of specific enteric pathogens from diarrheal data. Because biases from the OR and RR methods are likely greatest in such settings with high diarrhea burden, these two methods may be sufficient to guide public health planning across a wide range of settings. In settings with low diarrhea burden, estimates from the OR and RR methods are likely be even less biased.

## Data Availability

The data are publicly available at: https://clinepidb.org/ce/app/workspace/analyses/DS_5c41b87221/new.

## Acknowledgement

None.

## Notes

Conflict of interest: We declare no conflict of interest

Sources of funding: This work was supported by grant INV 065751 (to ERM and BL) from the Gates Foundation

### Competing Interest Statement

The authors have declared no competing interest.

### Funding Statement

This work was supported by grant INV 065751 (to ERM and BL) from the Gates Foundation

### Author Declarations

The study used ONLY openly available human data that were originally located at: https://clinepidb.org/ce/app/workspace/analyses/DS_5c41b87221/new.

## References

1. Lanata CF, Fischer-Walker CL, Olascoaga AC, Torres CX, Aryee MJ, Black RE. Global Causes of Diarrheal Disease Mortality in Children <5 Years of Age: A Systematic Review. PLoS One [Internet]. 2013 Sep 4 [cited 2024 Dec 10];8(9):e72788. Available from: https://journals.plos.org/plosone/article?id=10.1371/journal.pone.0072788

2. Lopman BA, Steele D, Kirkwood CD, Parashar UD. The Vast and Varied Global Burden of Norovirus: Prospects for Prevention and Control. PLoS Med [Internet]. 2016 Apr 1 [cited 2024 Dec 10];13(4):e1001999. Available from: https://journals.plos.org/plosmedicine/article?id=10.1371/journal.pmed.1001999

3. Kirk MD, Pires SM, Black RE, Caipo M, Crump JA, Devleesschauwer B, et al. World Health Organization Estimates of the Global and Regional Disease Burden of 22 Foodborne Bacterial, Protozoal, and Viral Diseases, 2010: A Data Synthesis. PLoS Med [Internet]. 2015 [cited 2025 Apr 21];12(12). Available from: https://pubmed.ncbi.nlm.nih.gov/26633831/

4. Grimprel E, Rodrigo C, Desselberger U. Rotavirus disease: Impact of coinfections. Pediatric Infectious Disease Journal [Internet]. 2008 Jan [cited 2024 Dec 16];27(1 SUPPL.). Available from: https://journals.lww.com/pidj/fulltext/2008/01001/rotavirus_diseaseimpact_of_coinfections.2aspx.

5. Troeger C, Blacker BF, Khalil IA, Rao PC, Cao S, Zimsen SR, et al. Estimates of the global, regional, and national morbidity, mortality, and aetiologies of diarrhoea in 195 countries: a systematic analysis for the Global Burden of Disease Study 2016. Lancet Infect Dis [Internet]. 2018 Nov 1 [cited 2024 Dec 19];18(11):1211–28. Available from: http://www.thelancet.com/article/S1473309918303621/fulltext

6. Liu J, Platts-Mills JA, Juma J, Kabir F, Nkeze J, Okoi C, et al. Use of quantitative molecular diagnostic methods to identify causes of diarrhoea in children: a reanalysis of the GEMS case-control study. Lancet [Internet]. 2016 Sep 24 [cited 2025 Aug 20];388(10051):1291. Available from: https://pmc.ncbi.nlm.nih.gov/articles/PMC5471845/

7. Platts-Mills JA, Babji S, Bodhidatta L, Gratz J, Haque R, Havt A, et al. Pathogen-specific burdens of community diarrhoea in developing countries: a multisite birth cohort study (MAL-ED). Lancet Glob Health [Internet]. 2015 [cited 2024 Dec 19];3(9):e564–75. Available from: https://pubmed.ncbi.nlm.nih.gov/26202075/

8. Pires SM, Fischer-Walker CL, Lanata CF, Devleesschauwer B, Hall AJ, Kirk MD, et al. Aetiology-Specific Estimates of the Global and Regional Incidence and Mortality of Diarrhoeal Diseases Commonly Transmitted through Food. PLoS One [Internet]. 2015 Dec 1 [cited 2024 Dec 11];10(12). Available from: https://pubmed.ncbi.nlm.nih.gov/26632843/

9. Ahmed SM, Hall AJ, Robinson AE, Verhoef L, Premkumar P, Parashar UD, et al. Global prevalence of norovirus in cases of gastroenteritis: a systematic review and meta-analysis. Lancet Infect Dis. 2014;14(8):725–30.

10. Rouhani S, Peñataro Yori P, Paredes Olortegui M, Siguas Salas M, Rengifo Trigoso D, Mondal D, et al. Norovirus Infection and Acquired Immunity in 8 Countries: Results From the MAL-ED Study. Clin Infect Dis [Internet]. 2016 May 15 [cited 2024 Dec 16];62(10):1210–7. Available from: https://pubmed.ncbi.nlm.nih.gov/27013692/

11. Blackwelder WC, Biswas K, Wu Y, Kotloff KL, Farag TH, Nasrin D, et al. Statistical Methods in the Global Enteric Multicenter Study (GEMS). 2012;

12. Rogawski McQuade ET, Liu J, Kang G, Kosek MN, Lima AAM, Bessong PO, et al. Protection From Natural Immunity Against Enteric Infections and Etiology-Specific Diarrhea in a Longitudinal Birth Cohort. J Infect Dis [Internet]. 2020 Nov 9 [cited 2023 Nov 28];222(11):1858–68. Available from: 10.1093/infdis/jiaa031

13. Houpt E, Gratz J, Kosek M, Zaidi AKM, Qureshi S, Kang G, et al. Microbiologic methods utilized in the MAL-ED cohort study. Clinical Infectious Diseases. 2014 Nov 1;59:S225–32.

14. Miller M, Acosta AM, Chavez CB, Flores JT, Olotegui MP, Pinedo SR, et al. The MAL-ED study: a multinational and multidisciplinary approach to understand the relationship between enteric pathogens, malnutrition, gut physiology, physical growth, cognitive development, and immune responses in infants and children up to 2 years of age in resource-poor environments. Clin Infect Dis [Internet]. 2014 Nov 1 [cited 2024 Jun 19];59 Suppl 4:S193–206. Available from: https://pubmed.ncbi.nlm.nih.gov/25305287/

15. Rogawski ET, Liu J, Platts-Mills JA, Kabir F, Lertsethtakarn P, Siguas M, et al. Use of quantitative molecular diagnostic methods to investigate the effect of enteropathogen infections on linear growth in children in low-resource settings: longitudinal analysis of results from the MAL-ED cohort study. Lancet Glob Health. 2018 Dec 1;6(12):E1319–28.

16. Platts-Mills JA, Liu J, Rogawski ET, Kabir F, Lertsethtakarn P, Siguas M, et al. Use of quantitative molecular diagnostic methods to assess the aetiology, burden, and clinical characteristics of diarrhoea in children in low-resource settings: a reanalysis of the MAL-ED cohort study. Lancet Glob Health [Internet]. 2018 Dec 1 [cited 2024 Dec 19];6(12):e1309–18. Available from: https://pubmed.ncbi.nlm.nih.gov/30287127/

17. Simmons K, Gambhir M, Leon J, Lopman B. Duration of Immunity to Norovirus Gastroenteritis - Volume 19, Number 8—August 2013 - Emerging Infectious Diseases journal - CDC. Emerg Infect Dis [Internet]. 2013 Aug [cited 2024 Feb 1];19(8):1260–7. Available from: https://www.nc.cdc.gov/eid/article/19/8/13-0472_article

18. Matthews JE, Dickey BW, Miller RD, Felzer JR, Dawson BP, Lee AS, et al. The epidemiology of published norovirus outbreaks: A review of risk factors associated with attack rate and genogroup. Epidemiol Infect [Internet]. 2012 Jul [cited 2025 Sep 6];140(7):1161–72. Available from: https://pubmed.ncbi.nlm.nih.gov/22444943/

19. Rockhill B, Newman B, Weinberg C. Use and misuse of population attributable fractions. Am J Public Health [Internet]. 1998 [cited 2025 Jan 1];88(1):15–9. Available from: https://pubmed.ncbi.nlm.nih.gov/9584027/

20. R Core Team. R: A language and environment for statistical computing. [Internet]. Vienna, Austria: R Foundation for Statistical Computing; 2018 [cited 2025 Sep 8]. Available from: https://www.R-project.org/

21. Bhavnani D, Goldstick JE, Cevallos W, Trueba G, Eisenberg JNS. Synergistic Effects Between Rotavirus and Coinfecting Pathogens on Diarrheal Disease: Evidence from a Community-based Study in Northwestern Ecuador. Am J Epidemiol [Internet]. 2012 Sep 1 [cited 2025 Jun 16];176(5):387. Available from: https://pmc.ncbi.nlm.nih.gov/articles/PMC3499114/

22. Blackwelder WC, Biswas K, Wu Y, Kotloff KL, Farag TH, Nasrin D, et al. Statistical methods in the Global Enteric Multicenter Study (GEMS). Clinical Infectious Diseases [Internet]. 2012 Dec 15 [cited 2025 Jun 17];55(SUPPL. 4). Available from: https://pubmed.ncbi.nlm.nih.gov/23169937/

