## Supplementary material for "Biases in Attribution Methods for Norovirus and Rotavirus Diarrhea": Online Appendix

#### 1. Model Equations

The ordinary differential equations for the model structure are as follows, whose corresponding states are listed on the left side of the equations.

| States | Ordinary differential equations[$] |
|---|---|
| Susceptible to infection | $\frac{dS}{dt} = \mu * N - (\lambda_{pathogen} + \lambda_{other} + \mu) * S + \delta I_{other}$ |
| Symptomatic initial infection | $\frac{dI_s}{dt} = \lambda_{pathogen} * s_1 * S - (\mu + \rho_s + \lambda_{other}) * I_s$ |
| Post-symptomatic shedding | $\frac{dI_{ps}}{dt} = \rho_s * I_s - (\mu + \lambda_{other} + \rho_{ps}) * I_{ps}$ |
| Asymptomatic initial infection | $\frac{dI_a}{dt} = \lambda_{pathogen} * (1 - s_1) * S - (\mu + \rho_a + \lambda_{other}) * I_a$ |
| Symptomatic reinfection | $\frac{dI_{sr}}{dt} = \lambda_{pathogen} * s_2 * R - (\lambda_{other} + \mu + \rho_s) * I_{sr}$ |
| Post-symptomatic shedding after reinfection | $\frac{dI_{psr}}{dt} = \rho_s * I_{sr} - (\lambda_{other} + \mu + \rho_{ps}) * I_{psr}$ |
| Asymptomatic reinfections | $\frac{dI_{ar}}{dt} = \lambda_{pathogen} * (1 - s_2) * R - (\rho_a + \lambda_{other} + \mu) * I_{ar}$ |
| Recovery | $\frac{dR}{dt} = \rho_a * (I_a + I_{ar}) + \rho_{ps} * (I_{ps} + I_{psr}) + \delta I_{coinf} - (\lambda_{other} + \lambda_{pathogen} + \mu) * R$ |
| Infection by other pathogens | $\frac{dI_{other}}{dt} = \lambda_{other} * (S + R) - (\delta + \lambda_{pathogen} + \mu) * I_{other}$ |
| Coiwnfection | $\frac{dI_{coinf}}{dt} = \lambda_{pathogen} * I_{other} + \lambda_{other} * (I_s + I_a + I_{ps} + I_{sr} + I_{ar} + I_{psr}) - (\delta + \mu) * I_{coinf}$ |

[$] $\lambda_{pathogen}$ is the force of infection for the target pathogen. $\lambda_{other}$ is the force of infection for other pathogens; $N$ is the total population size in the model.

$\lambda_{pathogen}$ was modeled as:

$$\lambda_{pathogen} = \beta_{pathogen} \frac{I_s + I_{sr} + I_{coinf} + \varepsilon(I_a + I_{ps} + I_{ar} + I_{psr})}{N}$$

where $\beta_{pathogen}$ is the transmission coefficient of the target pathogen, and ε represents a factor of reduction in infectiousness of asymptomatically infected persons relative to those with symptomatic infection.

$\lambda_{other}$ was modeled as:

$$\lambda_{other} = \beta_{other} \frac{I_{other} + I_{coinf}}{N}$$

where $\beta_{other}$ is the transmission coefficient for the non-target pathogens. We assume the individuals who are coinfected with the target and non-target pathogens, or those infected with the non-target pathogens, contribute to $\lambda_{other}$.

### 2. Likelihood Definition

We have three pieces of information based on the data collection.

1) Observed number of diarrhea (symptomatic) events, denoted as $n_{sym}$, for a given number of person-years in the sample, denoted as $PY_{sym}$.

2) Out of the $n_{sym}$ diarrheal samples, $n_{sym,pathogen}$ were positive for the target pathogen. This piece of data is dependent on 1) because the testing for the target pathogen in individuals experiencing diarrhea is dependent on the occurrence of diarrhea.

3) Out of the frequency of non-diarrheal samples from asymptomatic (ASx) individuals without diarrhea ($n_{ASx}$), $n_{ASx,pathogen}$ were positive for the target pathogen. Because the sampling scheme for the asymptomatic samples was independent from the sampling scheme for the symptomatic samples in MAL-ED (1), 3) is independent of 1) and 2).

The joint probability of these three pieces of data P[$n_{sym} \cap n_{sym,pathogen} \cap n_{ASx,pathogen}$] can then be simplified to P[$n_{sym}$]*P[$n_{sym,pathogen} | n_{sym}$]*P[$n_{ASx,parhogen}$].

We assume that P[$n_{sym}$] follows a Poisson distribution with a mean equal to the modeled number of diarrhea events. This mean is given by the product of the modeled all-cause diarrhea incidence at equilibrium and $PY_{sym}$:

$$\mathrm{P}[n_{sym}] = \mathrm{Poisson}\left(x = n_{sym}, \mu = \left(365 * \frac{(I^*_{other} + I^*_{coinf}) * \delta + (I^*_s + I^*_{sr}) * \rho_s}{N}\right) * PY_{sym}\right)$$

We assume that P[$n_{sym,pathogen} | n_{sym}$] follows a binomial distribution, where $n_{sym,pathogen}$ of the $n_{sym}$ diarrheal samples test positive for the target pathogen. The modeled steady-state prevalence of the pathogen among diarrheal samples is given by $\frac{I^*_s + I^*_{sr} + I^*_{coinf}}{I^*_s + I^*_{sr} + I^*_{coinf} + I^*_{other}}$. Hence,

$$\mathrm{P}[n_{sym,pathogen} | n_{sym}] = \mathrm{Binom}\left(x = n_{sym,pathogen}, n = n_{sym}, p = \frac{I^*_s + I^*_{sr} + I^*_{coinf}}{I^*_s + I^*_{sr} + I^*_{coinf} + I^*_{other}}\right)$$

Finally, we assume that P[$n_{ASx,pathogen}$] follows a binomial distribution, where $n_{ASx,pathogen}$ of the $n_{ASx}$ non-diarrheal samples are positive for the target pathogen. The modeled steady-state prevalence of the pathogen in non-diarrheal samples is given by $\frac{I^*_a + I^*_{ar} + I^*_{ps} + I^*_{psr}}{I^*_a + I^*_{ar} + I^*_{ps} + I^*_{psr} + S^* + R^*}$. Hence,

$$P[n_{ASx,pathogen}] = \text{Binom}\left(x = n_{ASx,pathgoen}, n = n_{ASx}, p = \frac{I^*{}_a + I^*{}_{ar} + I^*{}_{ps} + I^*{}_{psr}}{I^*{}_a + I^*{}_{ar} + I^*{}_{ps} + I^*{}_{psr} + S^* + R^*}\right)$$

Therefore, our joint likelihood for estimating the transmission rates of the target pathogen ($\beta_{pathogen}$) and other pathogens ($\beta_{other}$) is the product of these three independent likelihoods:

$$L(\beta_{pathogen}, \beta_{other}, s_1, s_2 | n_{sym}, PY_{sym}, n_{sym,pathogen}, n_{ASx,pathogen}, n_{ASx}) = P[n_{sym}] * P[n_{sym,pathogen} | n_{sym}] * P[n_{ASx,pathogen}]$$

Table S1. Fixed parameters for compartmental model of natural history of enteric infection

| Parameter | Notation | Value for norovirus | Value for rotavirus | Source/interpretation based on MAL-ED TaqMan Array Card data |
|---|---|---|---|---|
| Duration (day) of symptomatic infection | $1/\rho_s$ | 5 | 5 | Durations of diarrhea, positive for norovirus GII/rotavirus, regardless of coinfection with other pathogens (2)[&] |
| Duration (day) of symptomatic infection by other pathogens | $1/\delta$ | 5 | 6 | Duration of diarrhea, negative for norovirus GII/rotavirus (2)[&] |
| Duration (day) of post-symptomatic shedding | $1/\rho_{ps}$ | 23 | 8.5 | McMurry, 2021 (3) |
| Duration (day) of asymptomatic infection | $1/\rho_a$ | 30 | 18 | Saito, 2014 (4) for norovirus<br>Mukhopadhya 2013 (5) for rotavirus |
| Relative infectiousness of asymptomatic infection vs. symptomatic infection | $\varepsilon$ | 0.05 | 0.1 | Simmons, 2013 (6) for norovirus<br>For rotavirus, we assumed that the asymptomatic population has higher infectiousness compared to that of norovirus. So, we set $\varepsilon$ to 0.1, reflecting a greater potential for asymptomatic transmission than is observed for norovirus |
| Per-capita birth and death rates | $\mu$ | 1/365 | 1/365 | We assumed a simple birth-death process, so the model remains demographically dynamic |

[&]Calculated from the MAL-ED TaqMan Array Card data

Table S2. Correlations between model-based population attributable fraction (MB PAF) and key epidemiologic outputs from the norovirus and rotavirus transmission models.

| | All-cause diarrhea incidence | | Coinfection prevalence in diarrheal cases | | Asymptomatic infection prevalence | |
|---|---|---|---|---|---|---|
| | Norovirus | Rotavirus | Norovirus | Rotavirus | Norovirus | Rotavirus |
| MB PAF | -0.81 (0.03)[&] | -0.09 (0.84) | -0.58 (0.17) | 0.54 (0.17) | -0.42 (0.35) | 0.62 (0.10) |

[&]Pearson correlation (p value)

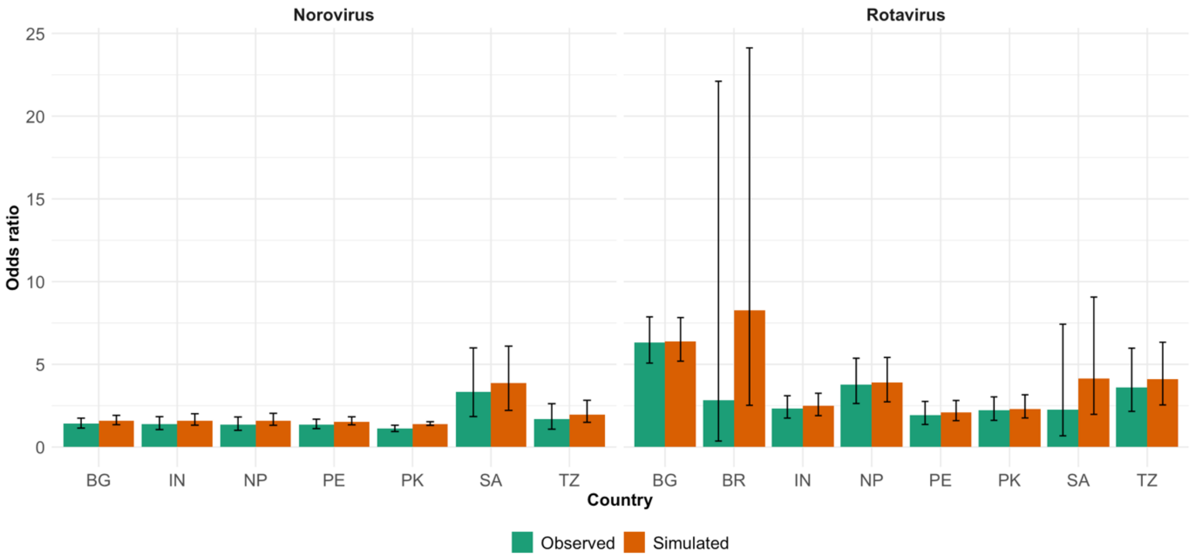


**Figure S1 Model-simulated site-specific odds ratios from the norovirus and rotavirus transmission models and their observed values.** Observed data are plotted with Wald-based 95% confidence intervals. Sites are labeled by country abbreviation: Bangladesh (BG), Brazil (BR), India (IN), Nepal (NP), Pakistan (PK), Peru (PE), South Africa (SA), Tanzania (TZ).

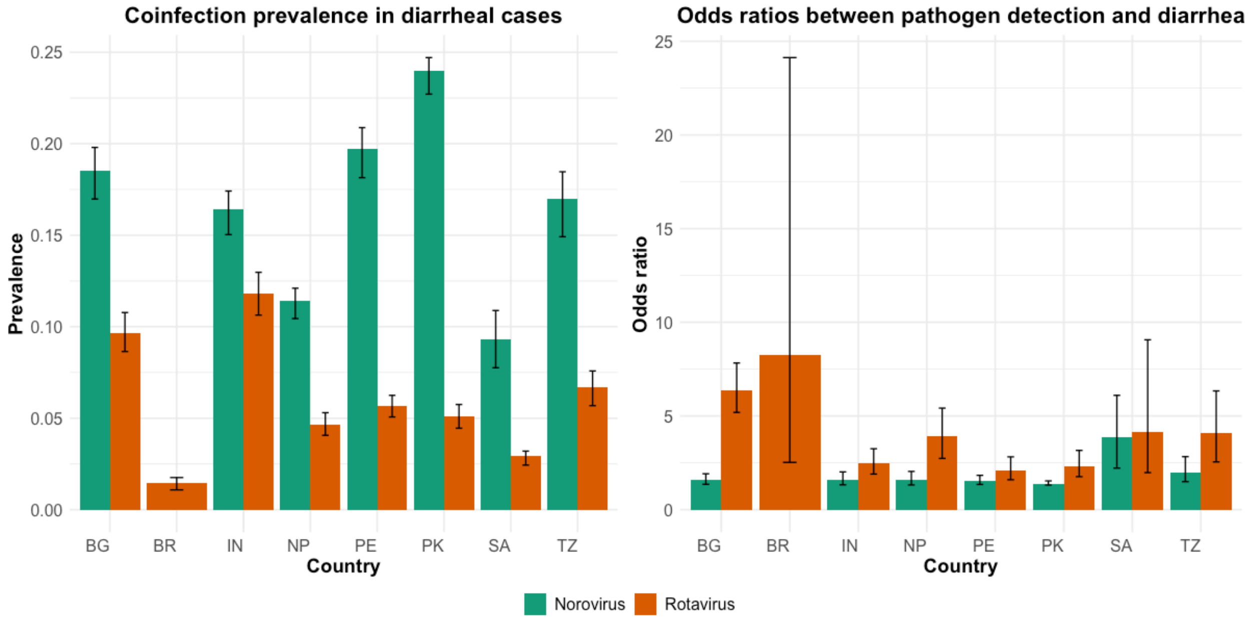


**Figure S2 Model-simulated site-specific symptomatic coinfection prevalence and odds ratio of pathogen detection and diarrhea from the norovirus and rotavirus models.** Sites are labeled by country abbreviation: Bangladesh (BG), Brazil (BR), India (IN), Nepal (NP), Pakistan (PK), Peru (PE), South Africa (SA), Tanzania (TZ).

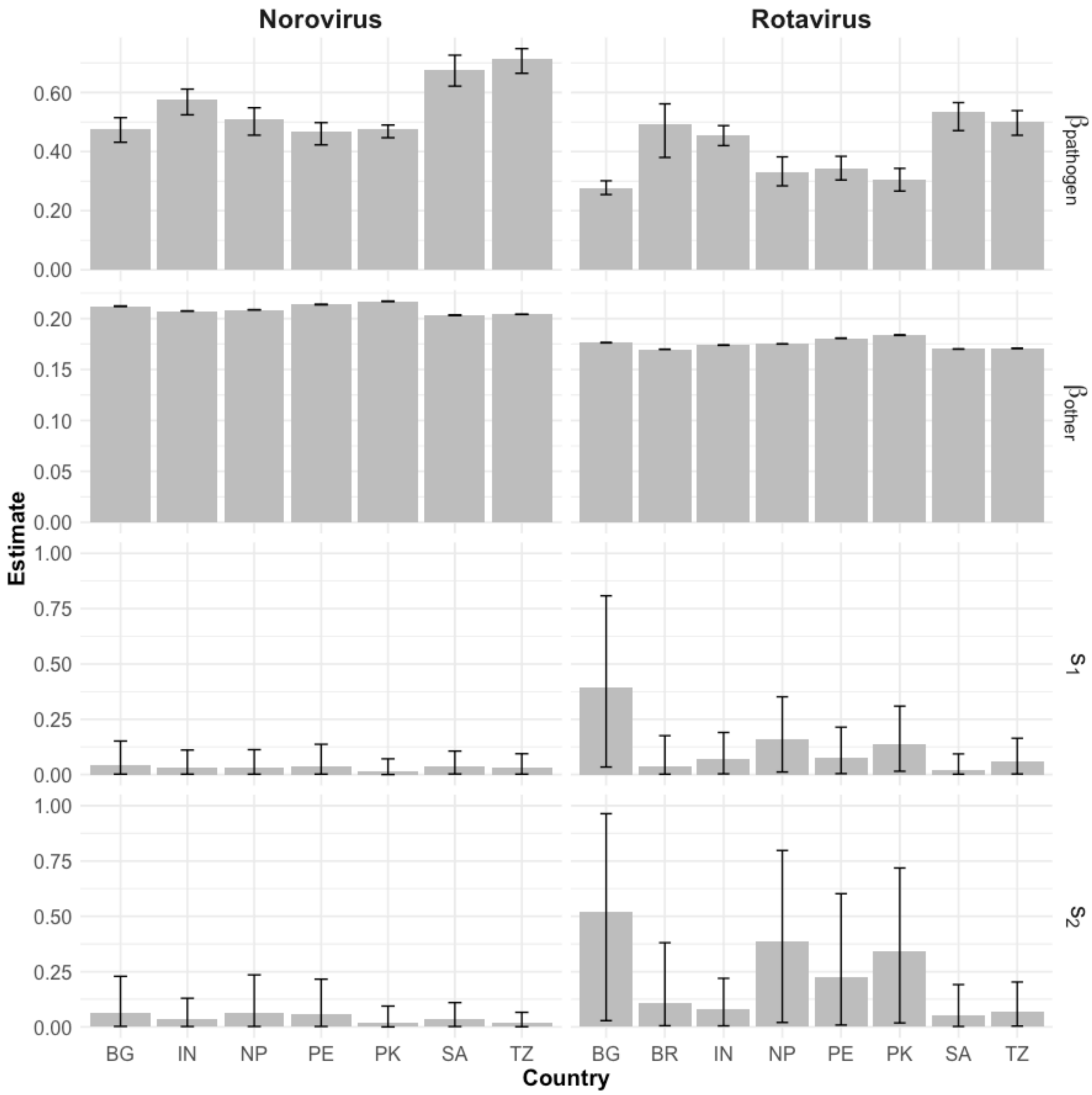


**Figure S3 Model-simulated site-specific parameters from the norovirus and rotavirus transmission models.** Model parameters include transmission coefficients for the target pathogen ($\beta_{pathogen}$) and other pathogens ($\beta_{other}$), and the probabilities of developing primary ($s_1$) and secondary ($s_2$) symptomatic infection in each model. Sites are labeled by country abbreviation: Bangladesh (BG), Brazil (BR), India (IN), Nepal (NP), Pakistan (PK), Peru (PE), South Africa (SA), Tanzania (TZ).

References


1. Miller M, Acosta AM, Chavez CB, Flores JT, Olotegui MP, Pinedo SR, et al. The MAL-ED study: a multinational and multidisciplinary approach to understand the relationship between enteric pathogens, malnutrition, gut physiology, physical growth, cognitive development, and immune responses in infants and children up to 2 years of age in resource-poor environments. Clin Infect Dis [Internet]. 2014 Nov 1 [cited 2024 Jun 19];59 Suppl 4:S193–206. Available from: https://pubmed.ncbi.nlm.nih.gov/25305287/

2. Spiro D, Gottlieb M, Glass R. Dataset: MAL-ED 0-60m Cohort [Internet]. 2022 [cited 2025 Jun 26]. Available from: https://clinepidb.org/ce/app/workspace/analyses/DS_5c41b87221/new

3. Mcmurry TL, Mcquade ETR, Liu J, Kang G, Kosek MN, Lima AAM, et al. Duration of Postdiarrheal Enteric Pathogen Carriage in Young Children in Low-resource Settings. Clinical Infectious Diseases [Internet]. 2021 Jun 1 [cited 2024 Feb 1];72(11):e806–14. Available from: https://dx.doi.org/10.1093/cid/ciaa1528

4. Saito M, Goel-Apaza S, Espetia S, Velasquez D, Cabrera L, Loli S, et al. Multiple Norovirus Infections in a Birth Cohort in a Peruvian Periurban Community. Clin Infect Dis [Internet]. 2014 Feb 2 [cited 2023 Nov 27];58(4):483. Available from: /pmc/articles/PMC3905757/

5. Mukhopadhya I, Sarkar R, Menon VK, Babji S, Paul A, Rajendran P, et al. Rotavirus shedding in symptomatic and asymptomatic children using reverse transcription-quantitative PCR. J Med Virol [Internet]. 2013 Sep [cited 2025 May 15];85(9):1661–8. Available from: https://pubmed.ncbi.nlm.nih.gov/23775335/

6. Simmons K, Gambhir M, Leon J, Lopman B. Duration of Immunity to Norovirus Gastroenteritis - Volume 19, Number 8—August 2013 - Emerging Infectious Diseases journal - CDC. Emerg Infect Dis [Internet]. 2013 Aug [cited 2024 Feb 1];19(8):1260–7. Available from: https://wwwnc.cdc.gov/eid/article/19/8/13-0472_article
